# Impact of disruptions in antigen processing and presentation machinery on sarcoma

**DOI:** 10.1101/2023.12.01.23299064

**Authors:** Salvatore Lorenzo Renne, Laura Sama’, Sonia Kumar, Omer Mintemur, Laura Ruspi, Ilaria Santori, Federico Sicoli, Alexia Bertuzzi, Alice Laffi, Arturo Bonometti, Piergiuseppe Colombo, Vittoria D’amato, Alessandra Bressan, Marta Scorsetti, Luigi Terracciano, Pierina Navarria, Maurizio D’incalci, Vittorio Quagliuolo, Fabio Pasqualini, Fabio Grizzi, Ferdinando Carlo Maria Cananzi

## Abstract

**Background:** The antigen processing machinery (APM) plays a critical role in generating tumor-specific antigens that can be recognized and targeted by the immune system. The status of the APM in sarcomas is not well characterized.

**Methods:** We investigated 126 patients with 8 types of bone and soft tissue sarcoma operated between 2001-2021. Tissue microarrays mapped 11 specific areas in each case. The presence/absence of APM protein was determined through immunohistochemistry. Bayesian networks were used.

**Results:** All investigated sarcomas had some defects in APM. The least damaged component was HLA Class I subunit β2-microglobulin and HLA Class II. The proteasome LMP10 subunit was defective in leiomyosarcoma (LMS), myxoid liposarcoma (MLPS), and dedifferentiated liposarcoma (DDLPS), while MHC I transporting unit TAP2 was altered in undifferentiated pleomorphic sarcoma (UPS), gastrointestinal stromal tumor (GIST), and chordoma (CH). Among different neoplastic areas, high-grade areas showed different patterns of expression compared to high lymphocytic infiltrate areas. Heterogeneity at the patient level was also observed. Loss of any APM component was prognostic of distant metastasis (DM) for LMS and DDLPS and of overall survival (OS) for LMS.

**Conclusion:** Sarcomas exhibit a high degree of defects in APM components, with differences among histotypes and tumoral areas. The most commonly altered APM components were HLA Class I subunit β2-microglobulin, HLA Class I subunit α (HC10), and MHC I transporting unit TAP2. The loss of APM components was prognostic of DM and OS and clinically relevant for LMS and DDLPS. This study explores sarcoma molecular mechanisms, enriching personalized therapeutic approaches.

## Introduction

Bone and soft tissue sarcomas are a heterogeneous group of rare tumors, comprising more than 80 histotypes, with different clinical behavior.^1^ Due to the great heterogeneity as regards histopathology, anatomic site and biologic potential, sarcomas are frequently diagnosed at advanced stages, with locally aggressive large tumors or metastatic disease. To the date, complete surgical resection remains the unique chance of cure. However, radically resected patients will fail in about 50% of cases, due to local recurrences or distant metastases.^2,3^ The addition of adjuvant chemotherapy didn’t show a significant improve of sarcoma patients outcomes.^4,5^

In the last years, immunotherapy emerged as an effective treatment for refractory malignant tumors.^6,7^ In this context, classifications based on the tumor micro-environment (TME) including of both the composition of the tumoral immune infiltrate and the expression of immune checkpoint ligands and receptors, was demonstrated significantly predictive and prognostic.^8^ However, limited evidences about sarcoma exist: a recent study from The Cancer Genome Atlas (TCGA) consortium suggested the association between the composition of sarcoma microenvironment (SME) and oncological prognosis.^9^ Nevertheless, only few reports investigating SME have been published.^10–12^ On the other hand efficacy of immunotherapy in sarcoma is yet to be defined.^13^

Antigen processing and presentation is necessary for an effective cytotoxic T cell activation and an intact antigen processing machinery (APM) is crucial for cancer control. Malignant transformation of cells is associated with down-regulation of HLA class I components in most of the tumors.^14^ After antigen release, uptake and presentation, primed and activated T cells infiltrate the tumor and thanks to the antigen presentation by HLA class I can recognize and kill the tumor cells.^15^ To present the antigen a tumor cell needs to have a proficient immunoproteasome, a proficient transporting protein and an intact Histocompatibility Complex class I (MHC I) composed by the α and the β subunits.^16^

The present study investigates the APM status in sarcoma.

## Materials and methods

### Patient selection

We explored 8 bone and soft tissue sarcoma histotypes: chordoma (CH), chondrosarcoma (CHS), gastrointestinal stromal tumor (GIST), retroperitoneal dedifferentiated liposarcoma (DDLPS), leiomyosarcoma (LMS), myxoid liposarcoma (MLPS), myxofibrosarcoma (MFS), undifferentiated pleomorphic sarcoma (UPS) and selected 126 patients (15 for each histotype and 21 for GIST) who underwent surgery at IRCCS Humanitas Research Hospital between 2001 and 2021. Diagnosis were made by sarcoma-expert pathologists and confirmed to fulfill the latest diagnostic criteria.^1^ Inclusion criteria were primary disease, availability of sufficiently abundant histologic material (see next section), and updated clinical follow-up. Metastases and local recurrences were excluded. The following data were extracted from our institutional prospectively maintained database: patient and tumor characteristics, administration of neoadjuvant or adjuvant treatments, type of surgery, vital status, and occurrence of local relapse and/or distant metastasis. Patient characteristics are summarized in supplementary table 1. The institutional review board of our hospital approved this retrospective study (145/23). Dataset is available on https://zenodo.org/communities/humanitasirccs.

### Tissue microarray construction

We built tissue microarray specifically for microenvironment characterization. We reviewed the slides, confirmed the diagnosis and selected 11 areas of each of the 126 cases: 3 high grade areas (defined as portions showing the highest mitotic activity, or cytologic atypia/pleomorphism); 1 low grade area (portion showing the lowest mitotic activity, or cytologic atypia/pleomorphism); 2 high infiltrate areas (portions with high lymphocytic infiltrate); 2 tumor periphery areas (tumoral portions within 1 mm from tumor-healthy tissue interface); 2 tumor border areas (portions of non-neoplastic tissue within 1 mm from healthy tissue-tumor interface); 1 non neoplastic area: portion of tissue far from the tumor. The presence of tumor heterogeneity (i.e. presence of high and low-grade areas) and of high infiltrate was also recorded for each case; if they were absent, we sampled additional high grade areas. TMA assembly was performed with an automated arrayer (TMA Grand Master - 3D HISTECH, Budapest, Hungary) using cores with a diameter of 1.0 mm.

### APM score determination

Formalin-fixed, paraffin-embedded TMA sections were treated with 3% hydrogen peroxide solution for 10 minutes at room temperature (RT) to quench endogenous peroxidase. Sections were then blocked with Background Sniper (Biocare Medicals, USA) for 20 min at RT and incubated with antibodies (clones) (see also supplementary table 2 for dilution) against proteasome component LMP10 (TO-7)^17^, MHC I transporting unit TAP2 (NOB2)^18^, HLA Class I subunit β2-microglobulin (SP0936)^17^, HLA Class I subunit α (HC10)^17^, HLA Class II (DPDQ)^19^. All the antibodies were kindly provided by Prof. Soldano Ferrone (Harvard Medical School, Boston, MA, United States). Slides were then incubated with MACH1 HRP-polymer for 30 min at RT (Biocare Medicals, USA). The chromogenic reaction was developed using 3,3’-Diaminobenzidine-tetrahydrochloride (DAB) Chromogen-Kit (Biocare Medicals, USA) and slides were counterstained with hematoxylin. For each TMA’s core, we recorded the status of the APM protein, as absent if there was total absence of staining on neoplastic cells, present if otherwise.

### Bayesian statistical analysis

Interactions of variables were modelled with Bayesian networks^20^ (Supplementary Figures 1 and 2). Models were fit using Stan (a probabilistic programming language) and R.^21,22^ CI was calculated as 89% of the highest posterior density interval. For survival modeling, APM was coded as *present* when the mean value of the *high-grade* cores was ≥ 0.6. Custom code showing model assumptions, model testing and diagnostics is available at https://github.com/slrenne/APM_Sarcoma.

## Results

### APM presence among histotypes

All the investigated sarcoma had some APM defects (Figure 1[post_pred_sim_APM]), the least damaged component being HLA Class I subunit β2-microglobulin and HLA Class II. Proteasome LMP10 subunit was likely to be defective in LMS, MLPS, and DDLPS with a mean posterior probability of 0.69 (CI: 0.28 – 1.00), 0.75 (CI: 0.60 – 0.92), and 0.83 (CI: 0.55 – 1.00) respectively. MHC I transporting unit TAP2 was likely to be altered in UPS, GIST, and CH with a mean posterior probability of 0.76 (CI: 0.39 – 1.00), 0.74 (CI: 0.55 – 0.94), and 0.84 (CI: 0.68 – 0.99) respectively. HLA Class I subunit β2-microglobulin was mostly lost in LMS, MLPS and DDLPS with a mean posterior probability density of 0.83 (CI: 0.53 –1.00), 0.85 (CI: 0.75 – 0.96), and 0.90 (CI: 0.70 – 1.00), respectively. HLA Class I subunit α (HC10) was instead lost in CHS, DDLPS and MFS with a mean posterior probability density of 0.68 (CI: 0.38 – 0.97), 0.81 (CI: 0.49 – 1.00), and 0.90 (0.68 – 1.00). Lastly HLA Class II was lost almost exclusively in MFS with a mean posterior probability density of 0.78 (CI: 0.32 – 1.00).

**Figure 1.**
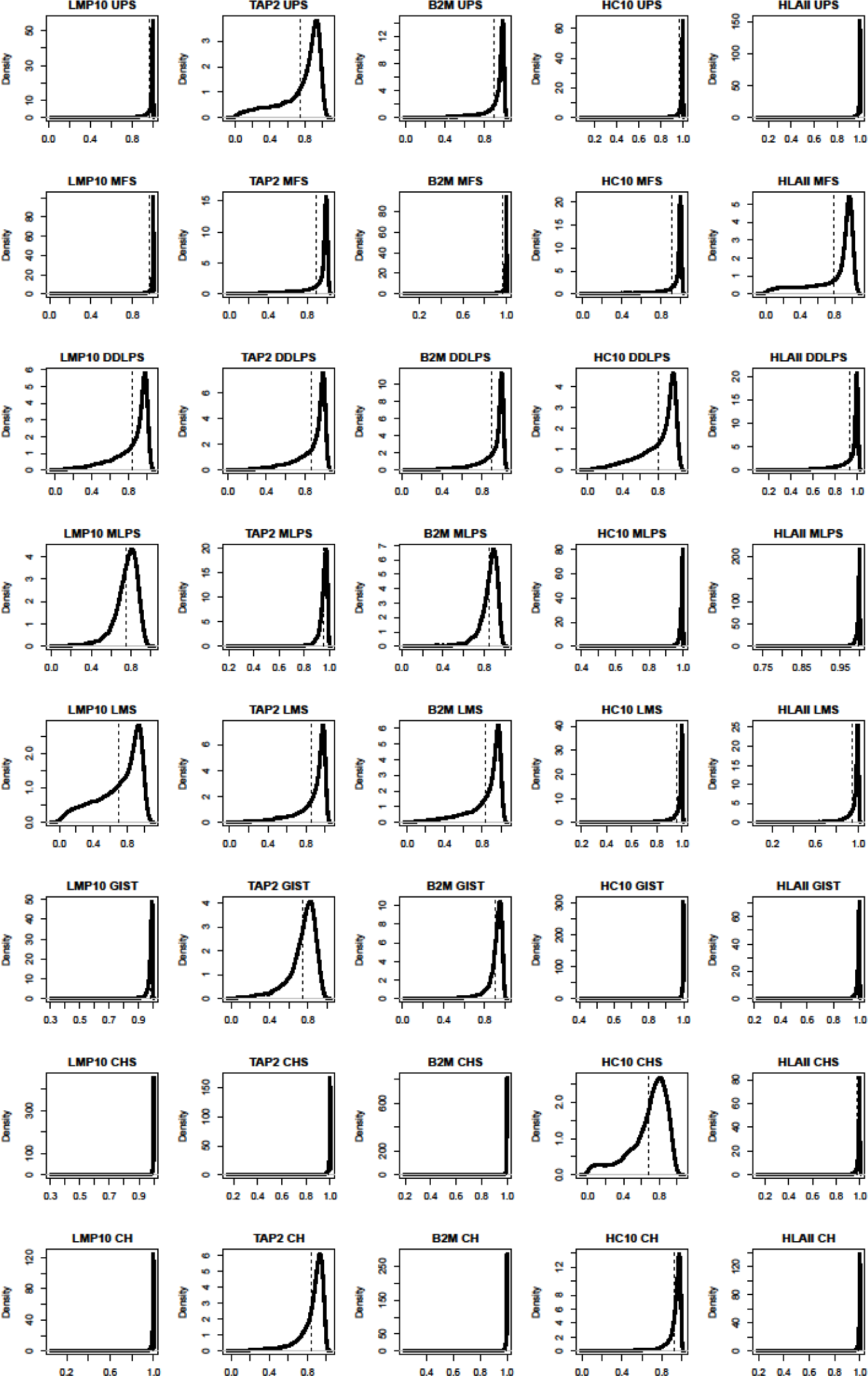
Probability of APM’s presence among sarcoma histotypes. Rows are the histotype and columns are the APM protein evaluated. Each figure shows the density (solid lines) of the probability of finding APM protein and the mean value (dashed vertical lines). LMP10 in UPS has most of the probability mass (i.e. the density) – as well as the mean – over 0.95; it is hence very unlikely to find defects in LMP10 in UPS. On the contrary, LMP10 in LMS can be defective in a substantial fraction of cases; similarly, MHC I transporting unit TAP2 was likely to be defective in UPS, GIST and HLA Class I subunit α (HC10) in CHS.

### Spatial differences of APM among tumoral areas and histotypes

We then investigated the different patterns of expression of APM in the different neoplastic areas (i.e. high grade, low grade, high infiltrate, tumor periphery, and tumor border) of each histotype (Figure 2[post_pred_sim_core]). These highlighted several differences, in particular between the areas of high grade compared with the areas of high infiltrate (Figure 3 [contrast_HGvsHI]). DDLPS cores with high infiltrate had less probability to lose proteasome LMP10 subunit expression compared to high grade areas with a mean contrast posterior probability density of -0.04 (CI: -0.08 – 0.00). Similarly, MHC I transporting unit TAP2 was likely to be retained in the high infiltrate areas of UPS and GIST compared to the high grade cores, with a mean contrast posterior probability density of -0.15 (CI: -0.26 – - 0.04) and -0.15 (-0.29 – -0.01) respectively. GIST also showed a similar trend with HLA Class I subunit β2-microglobulin expression having a greater probability of its loss in the high grade areas, with a mean contrast posterior probability density of -0.06 (-0.12 – 0.00). Lastly, HLA Class I subunit α (HC10) was more likely to be retained in the high infiltrate areas of CHS with a mean contrast posterior probability density of -0.04 (-0.09 – 0.00).

**Figure 2.**
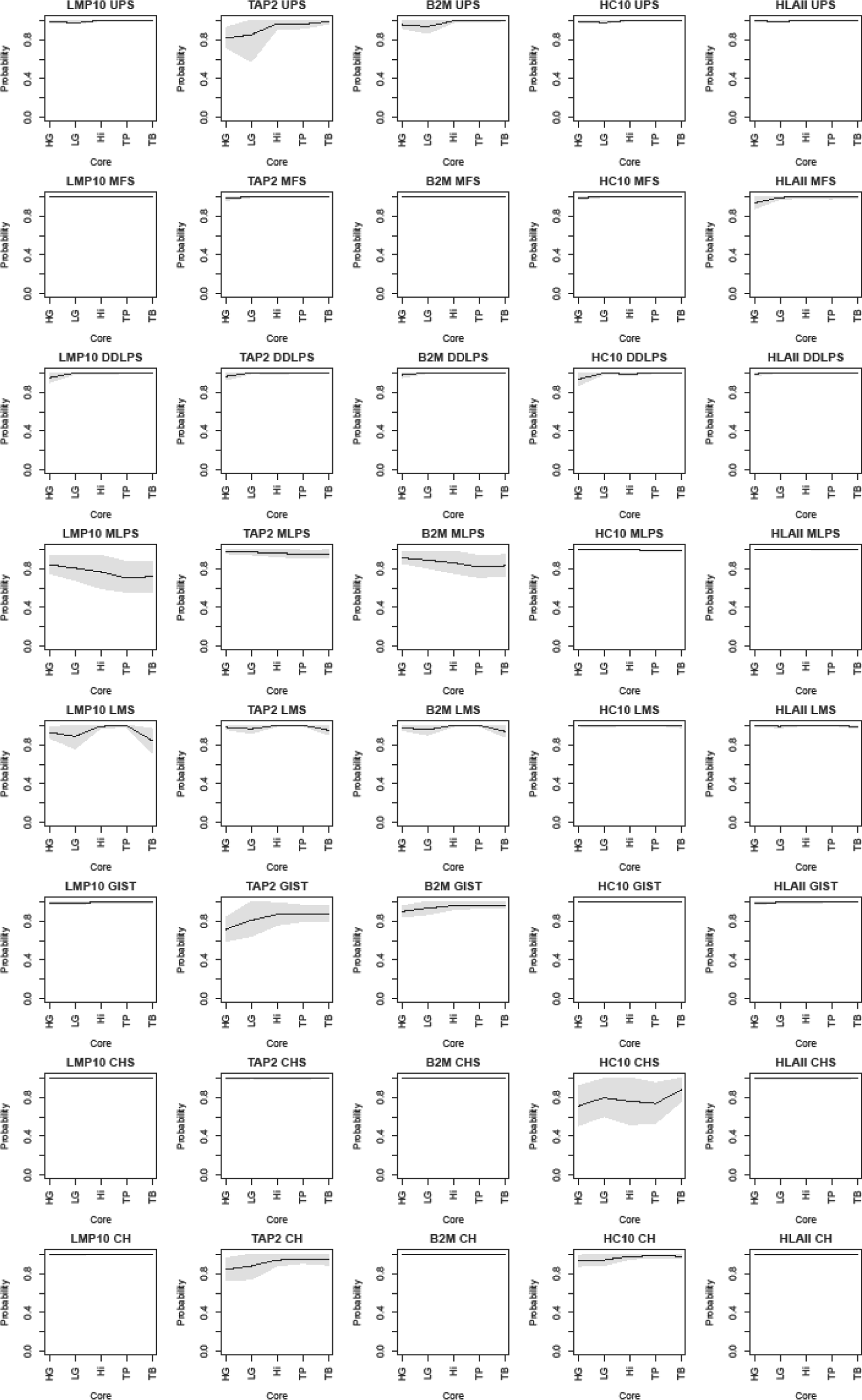
Spatial heterogeneity in the probability of APM’s presence among sarcoma histotypes. Rows are the histotype and columns are the APM protein evaluated. Each figure shows the mean (line) and CI (shade) for each ‘core-type’ evaluated. HG: high grade; HI: high infiltrate; LG: Low grade; HI: High infiltrate; TP: Tumor periphery, TB: Tumor border. See text for further details.

**Figure 3.**
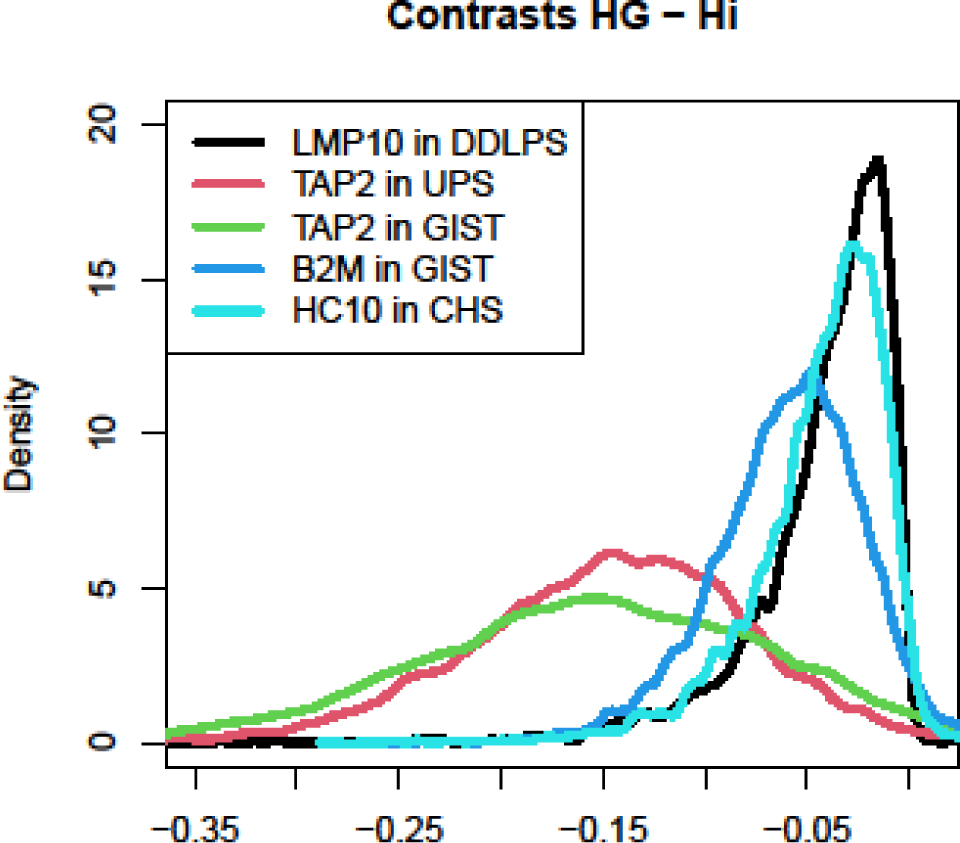
GIST, DDLPS, UPS, and CHS lose APM in high-grade areas. We computed the contrast with the High-Infiltrate areas. In fact, the latter are defined as those areas with high lymphocytic infiltrate, therefore where APM function is somehow preserved. The densities of the contrast show that almost all the probability mass fall below 0, indicating a substantial decrease in the probability of having that peculiar APM protein in that specific histotype.

### Interpatients variability of APM among tumoral areas and histotypes

We therefore investigated the heterogeneity at patient level. To do so we draw 10 random samples from the posterior probability distribution for each combination of the APM subunit and the core type (Figure 4 [Post_Pred_Sim_APM_core_10ptv2]). This analysis showed some patient was expected to have more often deficiency of some particular APM component in a specific core: as in the cases of the MHC I transporting unit TAP2 in UPS and HLA Class I subunit β2-microglobulin in CH. However, most of the effect was uniform within each histotype reflecting a lower interpatient variability once the histotype and the type of tumoral area were known.

**Figure 4.**
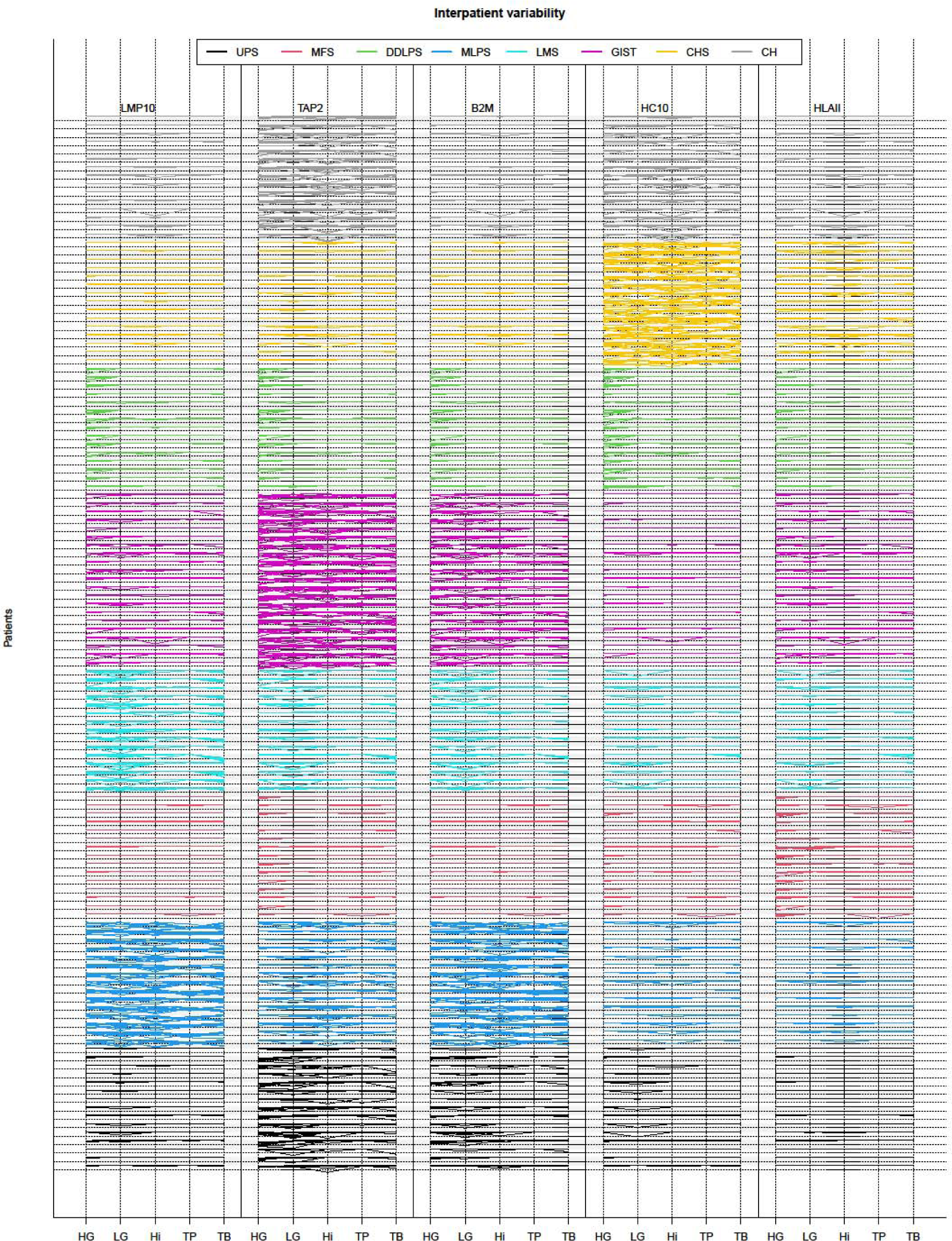
Interpatient variability. The figure shows 10 realizations for each patient, for each TMA core, and for each APM protein (i.e. the model representation of the data, with the advantage to show variability). Each column shows all the 126 patients, where the histotype is highlighted by the color and the core by the bending sites. The five columns are the APM protein tested. Cases that show greater uncertainty show more dispersed lines.

### Survival Analysis

We finally investigated the role of APM component loss in each histotype both for distant metastases and overall survival (Figure 5 [DFS&OS]). Given the assumptions depicted in the directed acyclic graph underlying the model (Supplementary Figure 1[APM_DM], 2[APM_OS]) to estimate the effect of the APM loss on distant metastases free survival and of overall survival we need to adjust for tumor grade. Loss of any of the APM component was prognostic of distant metastasis for LMS and DDLPS with mean hazard ratio of 26.57 (CI: 2.39 – 86.17) and 9.75 (CI: 1.03 – 31.04) respectively (Supplementary Figure 3 [Post_DFS_HR_g]). Similarly, loss of any of the APM component was prognostic of Overall Survival for LMS and DDLPS with mean hazard ratio of 86.15 (CI: 1.06 – 87.86) and 2.91 (CI: 0.39 – 8.32) respectively (Supplementary Figure 4 [Post_OS_HR_g]).

**Figure 5.**
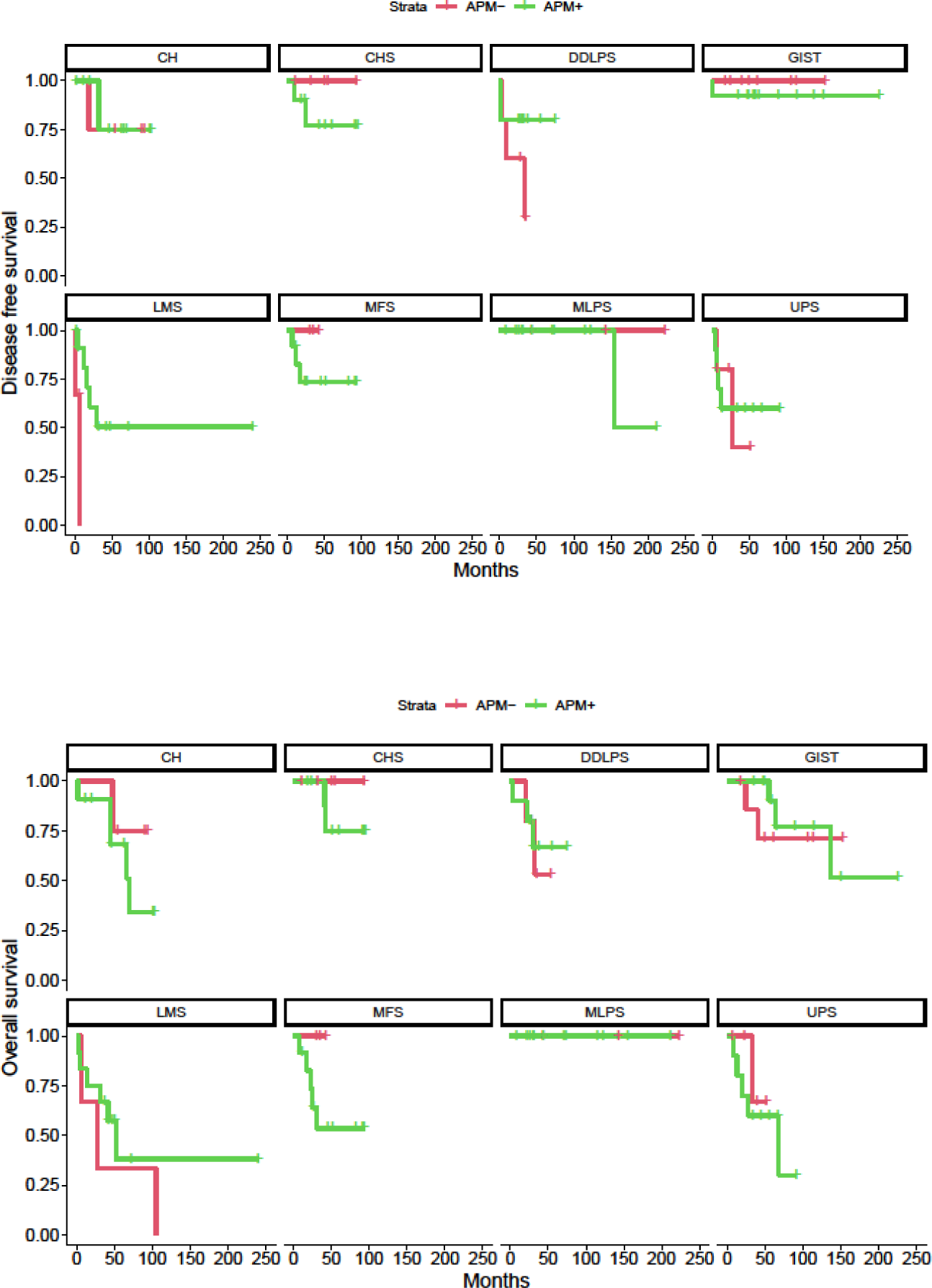
**Kaplan-Meier plots for disease free survival (DFS) and overall survival (OS) in APM proficient and deficient tumors.**

## Discussion

We here characterized with immunohistochemistry the main components of the antigen processing and presenting machinery (APM), specifically studying the proteasome subunit LMP10, the MHC I transporting unit TAP2, the HLA Class I subunit β2-microglobulin, the HLA Class I subunit α, and the HLA Class II in a cohort of 126 cases This is the first study that systematically study the APM in sarcoma and showed the difference of APM presence among the most prevalent histotypes, we explored the spatial and interpatients difference, and finally the prognostic value of APM loss.

The first attempt to highlight the role of APM in sarcoma was pioneered by Tsukahara et al. that characterized HLA Class 1 ABC and β2-microglobulin in a large series of bone (comprising 33 osteosarcomas and 5 chondrosarcomas) and soft tissue tumors (among them 15 undifferentiated pleomorphic sarcomas, 5 liposarcomas, 3 rhabdomyosarcomas, and 2 cases of Ewing, clear cell, synovial, alveolar soft part, leiomyosarcoma and 1 malignant peripheral nerve sheath tumor.^23^ In this seminal work, they found a prognostic role for APM loss in osteosarcoma, but not in UPS – finding confirmed in our analysis – however their cohort of osteosarcomas comprised both primary and metastatic diseases. From the same group Yabe et al. studied the same molecules in 28 Ewing sarcoma family cases where they found a prognostic role for APM.^24^ In contrast with these publications, we focused on the most prevalent sarcomas.^25^ Moreover, we mapped the neoplasm including different areas from multiple blocks with 11 cores for each case. Tissue microarrays constructed punching multiple selected areas from a single tumor are comparable for tissue description to whole-slide.^26^ Given the extensive mapping of our cases, TMA might be even more effective than whole slide staining in sarcoma given the size and higher degree of heterogeneity compared to other (non-mesenchymal) tumor types.

These differences with other tumor types emerged also in a lower rate of defect in all the components of APM.^14^ Indeed, size and heterogeneity pose an important challenge in studying sarcoma and microenvironment; however, we mapped the tumor heterogeneity and this enable us to demonstrate that within the same tumor the areas with inflammatory cells had a preserved APM expression compared to the high grade area, that is the one that drive the prognosis.^27^ In particular, this was true for LMP10 in DDLPS, TAP2 in UPS and GIST, β2-microglobulin in GIST, and HLA Class I subunit α in CHS.

We also investigated the heterogeneity at patient level; indeed, our statistical model was able to gather the difference among each single patient using a multilevel structure.^28^ This approach allowed us to see that most of the differences occurred among histotypes and the element of the APM studied, and that the differences of patients within these two categories were minimal. This finding may be reassuring and underlines the importance of a histological diagnosis, relevance that has been underlined also by genomic data.^9^

Lastly, similar to many other histotypes, we found APM loss associated with a worse distant metastases free survival and overall survival in LMS and DDLPS.^14^ This is expected because the generation of the immune response against cancer is – also – due to proficient APM,^15^ and therefore the absence of APM will lead, even in the presence of correctly primed cytotoxic T-cells, in absence in cancer recognition and consequently tumor survival.^16^ On the other hand the absence of association with survival cannot be interpreted as the evidence that this alteration are not relevant in the other cancer types (namely UPS, MFS, MLPS, GIST, CHS, and CH), indeed APM loss represents just one of the way in which cancer can avoid immune destruction,^15^ and this avoidance is just one small facet of the mechanisms that cancer need to acquire to thrive.^29,30^

The study cohort was limited to a relatively small sample size of 126 cases. A larger sample size would provide more robust data however we employed multilevel hierarchical models that are the state of the art technique to draw inference and regularize even in presence of small sample size.^28^ Moreover, the study focused on a limited number of APM components. While the selected proteins are key players in the antigen processing and presentation pathway, other components, such as the chaperone proteins tapasin and calreticulin, were not examined. Future studies that examine a more comprehensive set of APM components would be useful. Finally, we did not examine the effect of APM dysregulation on immune cell infiltration or function within the tumor microenvironment. Future studies that examine the impact of APM dysregulation on the recruitment and function of different immune cell populations, such as T cells and natural killer cells, would provide valuable insights.

In summary, we here systematically characterized the expression of several APM proteins in a series of 126 soft tissue, viscera and bone sarcomas. This analysis revealed multiple APM defects among the histotypes, highlighting the heterogeneity and complexity of the tumor microenvironment. The study also demonstrated that when lymphocytes are infiltrating the tumor tissue, APM expression tends to be preserved, suggesting a potential role for immune cells in promoting an anti-tumor immune response. Furthermore, the study showed that APM defects were found to be prognostic in certain sarcoma types, indicating that the presence or absence of APM expression could be used as a marker for predicting patient outcomes. Overall, these findings help to elucidate the intricate interactions between tumor cells and the microenvironment and provide important insights for developing more effective strategies for the diagnosis and treatment of sarcomas. Future research will be focused on the validation of these findings in larger and more diverse cohorts of sarcoma patients to confirm the prognostic value of APM loss in DDLPS and LMS; the investigation of the underlying mechanisms responsible for APM loss in sarcoma, including the role of genetic alterations, epigenetic modifications, and the tumor microenvironment, and finally the evaluation of the impact of APM preservation on immune cell infiltration and function within the tumor microenvironment.

In conclusion, our study provides a comprehensive characterization of the antigen processing and presenting machinery (APM) in sarcomas, identifying differences in APM presence among the most prevalent histotypes, exploring the spatial and inter-patient differences, and revealing the prognostic value of APM loss in DDLPS and LMS. This work sheds light on the complexity of the tumor microenvironment and highlights the importance of incorporating APM status in sarcoma diagnosis and treatment. Our findings provide a basis for further research to develop targeted therapies and personalized treatment approaches for sarcoma patients.

## Data Availability

All data produced are available online at https://zenodo.org/communities/humanitasirccs.

## Acknowledgements

This study was supported by Ricerca Finalizzata 2021 by Italian Ministry of Health - Giovani Ricercatori (GR) - “Change promoting”, project code GR-2021-12373209. We express our gratitude to the reSeARChOMA group, the institutional translational research group on sarcoma of Humanitas University.

