## Supplementary materials for "Impact of disruptions in antigen processing and presentation machinery on sarcoma"

### Supplementary material

***Supplementary Table 1:*** *Patients Clinico-pathological characteristics*

| | |
|---|---|
| Age, median (Q1-Q3) | 63.4 (51.75-74.23) |
| Women, No. (%) | 65 (51.6) |
| Men, No. (%) | 61 (48.4) |
| Tumor Size (cm), median (Q1-Q3) | 8.25 (5.6-13.8) |
| Neoadjuvant treatment No. (%) | |
| Yes | 8 (6.3) |
| CT | 7 (5.5) |
| RT | 1 (0.8) |
| CT/RT | 0 |
| No | 118 (93.7) |
| Adjuvant treatment No. (%) | |
| Yes | 61 (48.4) |
| CT | 15 (11.9) |
| RT | 36 (28.6) |
| CT/RT | 5 (3.9) |
| No | 65 (51.6) |
| Resection margins, No. (%) | |
| R0 | 104 (82.5) |
| R1 | 18 (14.3) |
| R2 | 4 (3.2) |
| Tumor rupture, No. (%) | |
| Yes | 5 (4.0) |
| No | 121 (96.0) |
| Presence of high infiltrate, No. (%) | |
| Yes | 41 (32.6) |
| No | 85 (67.4) |
| Tumor heterogeneity, No. (%) | |
| Yes | 51 (40.5) |
| No | 75 (59.5) |
| Local recurrence (LR), No. (%) | |
| Yes | 31 (24.6) |
| No | 95 (75.4) |
| Time to LR (months), median (Q1-Q3) | 14 (9.9-22.25) |
| Distant metastasis (DM), No. (%) | |
| Yes | 28 (22.2) |
| No | 98 (77.8) |
| Time to DM (months), median (Q1-Q3) | 10.77 (5.35-21.1) |
| Patient status, No. (%) | |
| alive, NED | 75 (59.5) |
| alive with disease | 13 (10.4) |
| died for disease | 30 (9.5) |
| died for other cause | 8 (6.3) |

Q: quartile; NED: no evidence of disease

***Supplementary Table 2:*** *Antibodies, Clones, Species, and Dilutions*

| Antibody | Clone | Species | Dilution |
|---|---|---|---|
| **HLA Class I** | HC10 | Mouse | 1:50 |
| **LMP10** | TO-7 | Mouse | 1:200 |
| **HLA Class II** | DPDQ | Mouse | 1:50 |
| **β2microglobulin** | SP09-36 | Mouse | 1:500 |
| **TAP2** | NOB-2 | Mouse | 1:500 |

Utilizing a Bayesian Network as a predictive model, we constructed a Directed Acyclic Graph (DAG) incorporating relevant clinico-pathological variables.

***Supplementary Model 1***

$$score \sim \text{dbinom}(1, p)$$
$$Logit(score) = \gamma_{NPL} + \alpha_{CORE,NPL} + \beta_{ID,NPL} + \delta_{APM,NPL}$$
$$\begin{bmatrix} \alpha_{j,1} \\ \vdots \\ \alpha_{j,8} \end{bmatrix} \sim MVNormal\left( \begin{bmatrix} 0 \\ \vdots \\ 0 \end{bmatrix}, H_\alpha \right)$$
$$H_\alpha = \begin{pmatrix} \sigma_{\alpha 1} & 0 & \dots & 0 \\ 0 & \sigma_{\alpha 2} & \dots & 0 \\ \vdots & \vdots & \ddots & \vdots \\ 0 & 0 & \dots & \sigma_{\alpha 8} \end{pmatrix} R_\alpha \begin{pmatrix} \sigma_{\alpha 1} & 0 & \dots & 0 \\ 0 & \sigma_{\alpha 2} & \dots & 0 \\ \vdots & \vdots & \ddots & \vdots \\ 0 & 0 & \dots & \sigma_{\alpha 8} \end{pmatrix}$$
$$\begin{bmatrix} \beta_{j,1} \\ \vdots \\ \beta_{j,8} \end{bmatrix} \sim MVNormal\left( \begin{bmatrix} 0 \\ \vdots \\ 0 \end{bmatrix}, H_\beta \right)$$
$$H_\beta = \begin{pmatrix} \sigma_{\beta 1} & 0 & \dots & 0 \\ 0 & \sigma_{\beta 2} & \dots & 0 \\ \vdots & \vdots & \ddots & \vdots \\ 0 & 0 & \dots & \sigma_{\beta 8} \end{pmatrix} R_\beta \begin{pmatrix} \sigma_{\beta 1} & 0 & \dots & 0 \\ 0 & \sigma_{\beta 2} & \dots & 0 \\ \vdots & \vdots & \ddots & \vdots \\ 0 & 0 & \dots & \sigma_{\beta 8} \end{pmatrix}$$
$$\begin{bmatrix} \delta_{j,1} \\ \vdots \\ \delta_{j,8} \end{bmatrix} \sim MVNormal\left( \begin{bmatrix} 0 \\ \vdots \\ 0 \end{bmatrix}, H_\delta \right)$$
$$H_\delta = \begin{pmatrix} \sigma_{\delta 1} & 0 & \dots & 0 \\ 0 & \sigma_{\delta 2} & \dots & 0 \\ \vdots & \vdots & \ddots & \vdots \\ 0 & 0 & \dots & \sigma_{\delta 8} \end{pmatrix} R_\delta \begin{pmatrix} \sigma_{\delta 1} & 0 & \dots & 0 \\ 0 & \sigma_{\delta 2} & \dots & 0 \\ \vdots & \vdots & \ddots & \vdots \\ 0 & 0 & \dots & \sigma_{\delta 8} \end{pmatrix}$$
$$\gamma_{NPL} \sim Normal(0, 1)$$
$$R_\alpha \sim LKJcorr(2)$$
$$R_\beta \sim LKJcorr(2)$$
$$R_\delta \sim LKJcorr(2)$$
$$\sigma_{\alpha j} \sim Exponential(1) \text{ for } j = 1..8$$
$$\sigma_{\beta j} \sim Exponential(1) \text{ for } j = 1..8$$
$$\sigma_{\delta j} \sim Exponential(1) \text{ for } j = 1..8$$

We analyzed APM defects by modeling it as a binomial distribution which showed as *score*. The score depends on the combination of NPL (histotypes) and its interrelation with Core (HG, LG, HI, TP, and TB), Id (patient ID), and APM (protein). Overall network was modeled as hierarchical Bayesian model. The model estimates parameters for NPL, Core and NPL, patient ID and NPL and finally APM and NPL. Samples for NPL had a normal distribution that had 0 mean and variance of 1. Samples for this model except NPL comes from multivariate normal distribution. The mean of this distribution for each relation was set to 0 and covariance matrix of the distribution was determined by variance covariance matrix for each relation. There were 8 histotypes. Since 8 different histotypes were analyzed, the covariance matrix for those relationships had a shape of 8x8. R value, which determines the correlation factor for each matrix was sampled from the LKJcorr distribution, with a mild regularizing prior of 2.[2-5]

***Supplementary Model 2***

$$T_i \mid Survival_i = 1 \sim Exponential(\lambda_i)$$
$$T_i \mid Survival_i = 0 \sim ExponentialLCCDF(\lambda_i)$$
$$\lambda_i = 1/\mu_i$$
$$Exponential(\mu_i) = \alpha_{APM,HIST} + \beta_{GRADE,HIST}$$
$$\begin{bmatrix} \alpha_{j,1} \\ \vdots \\ \alpha_{j,8} \end{bmatrix} \sim MVNormal\left(\begin{bmatrix} 0 \\ \vdots \\ 0 \end{bmatrix}, H_\alpha\right)$$
$$H_\alpha = \begin{pmatrix} \sigma_{\alpha1} & 0 & \dots & 0 \\ 0 & \sigma_{\alpha2} & \dots & 0 \\ \vdots & \vdots & \ddots & \vdots \\ 0 & 0 & \dots & \sigma_{\alpha8} \end{pmatrix} R_\alpha \begin{pmatrix} \sigma_{\alpha1} & 0 & \dots & 0 \\ 0 & \sigma_{\alpha2} & \dots & 0 \\ \vdots & \vdots & \ddots & \vdots \\ 0 & 0 & \dots & \sigma_{\alpha8} \end{pmatrix}$$
$$\begin{bmatrix} \beta_{j,1} \\ \vdots \\ \beta_{j,8} \end{bmatrix} \sim MVNormal\left(\begin{bmatrix} 0 \\ \vdots \\ 0 \end{bmatrix}, H_\beta\right)$$
$$H_\beta = \begin{pmatrix} \sigma_{\beta1} & 0 & \dots & 0 \\ 0 & \sigma_{\beta2} & \dots & 0 \\ \vdots & \vdots & \ddots & \vdots \\ 0 & 0 & \dots & \sigma_{\beta8} \end{pmatrix} R_\beta \begin{pmatrix} \sigma_{\beta1} & 0 & \dots & 0 \\ 0 & \sigma_{\beta2} & \dots & 0 \\ \vdots & \vdots & \ddots & \vdots \\ 0 & 0 & \dots & \sigma_{\beta8} \end{pmatrix}$$
$$R_\alpha \sim LKJcorr(2)$$
$$R_\beta \sim LKJcorr(2)$$
$$\sigma_{\alpha j} \sim Exponential(1)_{\text{for } j = 1..8}$$
$$\sigma_{\beta j} \sim Exponential(1)_{\text{for } j = 1..8}$$

We performed a survival analysis modeling the time-to-event ($T_i$) as an exponential distribution and its log-complementary-cumulative-density-function, similarly to Cox survival analysis[1]. The expected rate ($\lambda_i$) was then converted to the expected value $\mu_i$ and used a log() as a link function to the linear model. It was an *intercept-only* model. $\alpha + \beta$ was modeled using a hierarchical Bayesian approach, estimating a parameter for the APM status and the histotype and for the grading and the histotype, with their parameters sampled from a multivariate normal distribution. The mean of this distribution was set to 0, and the covariance matrix was determined by variance-covariance matrix of the presence of APM (and the grade) and the 8 histotypes, denoted as H. Given that there were a total of 8 sampled histotypes, covariance matrix had a shape of 8x8. The correlation factor within the matrix was determined by R, which was sampled from the LKJcorr distribution with a chosen shape parameter of 2.[2-5]

### Supplementary Figure Legends

Supplementary Figure 1. **Minimal adjustment set to estimate the effect of APM on Disease Free Survival (DFS).** The DAG models the effect of APM on DFS. The adjustment set demand, conditioning on grade in each histotype. Blue circle shows the outcome of modeled Bayesian Network. The green circle denotes the APM variable that model assessed the effect of it.

Model was figured using a web page called daggity. (https://dagitty.net)

Supplementary Figure 2. **Minimal adjustment set to estimate the effect of APM on Overall Survival (OS).** The Bayesian Network model's DAG is more complicated than the first model since this model considers more independent variables. This model, similar to DFS model, assess the effects of APM presence in OS.

Model was figured using a web page called daggity. (https://dagitty.net)

Supplementary Figure 3. **Disease Free Survival Hazard Ratio Grade Included.** Each row presents an effect of APM loss on specific histotype. Overall, the figure shows the hazard ratio between APM existence and APM loss. LMS and DDLPS histotypes were affected severely from the APM loss in DFS.

Supplementary Figure 4. **Overall Survival Hazard Ratio Grade Included.** Each row shows the same information as mentioned in Supplementary Figure 3. Similarly, both LMS and DDLPS were the same histotypes that were affected from APM loss in OS.
